# “*We need to highlight as a community that these are avoidable*”: Exploring clinician perspectives on healthcare access and avoidable admissions in inflammatory bowel disease

**DOI:** 10.1101/2025.09.12.25335651

**Authors:** Rachel L Hawkins, Matthew Lee, Fiona Sampson, Daniel Hind, Alan J Lobo

**Affiliations:** Sheffield Centre for Health and Related Research, School of Medicine and Population Health, University of Sheffield, Sheffield, United Kingdom; Sheffield Inflammatory Bowel Disease Centre, Sheffield Teaching Hospitals NHS Foundation Trust, Sheffield, United Kingdom; School of Healthcare, The University of Leeds, Leeds, United Kingdom; Department of Applied Health Sciences, University of Birmingham, Birmingham, United Kingdom

## Abstract

**Objectives:** Unplanned hospital admissions are common for people with inflammatory bowel disease (IBD). While clinical predictors of admission are well-documented, research is limited exploring the healthcare delivery factors and system inequalities that contribute to these events.

**Design:** An online survey, distributed via United Kingdom (UK) and European professional networks, and semi-structured interviews explored clinicians’ views of admission types, causes and barriers to preventing admissions. Clinicians ranked causes and barriers to preventing admissions, analysed by descriptive statistics and Friedman and Wilcoxon rank tests. We used framework analysis, guided by Candidacy Theory, to analyse qualitative data across data types to explore issues of healthcare access.

**Results:** 80 clinicians completed the survey, 13 were interviewed. For all unplanned IBD admissions, an unpreventable disease progression was ranked the highest cause (mean rank [MR] 32.20/100) (*p* <0.05), followed by missed opportunities for earlier intervention (MR 22.39/100) and patient access issues (MR 19.52/100). Qualitative interviews elaborated on avoidable unplanned admissions through: (1) Missed opportunities in outpatient or primary care, (2) critical delays in specialised care, and (3) system constraints blocking timely action. Key challenges in preventing admissions across the study related to patient navigation of services, organisational barriers, provider decision-making and structural issues that impede access to care.

**Conclusions:** Most avoidable admissions were perceived not as a failure of individuals, but a reflection of broader service inefficiencies and inequities. Reducing IBD admissions requires systemic investment and improvements in care navigation, rapid-access pathways, professional decision-making, patient education and service integration.

**What is already known on this topic:**

⍰ Unplanned hospital admissions are a common and costly outcome for people living with inflammatory bowel disease (IBD).
⍰ While clinical predictors of admission are well-documented, there is limited research exploring the healthcare delivery factors and system inequalities that contribute to these events.

**What this study adds:**

⍰ This study presents a clinician-led definition of avoidable IBD admissions as those resulting from missed outpatient opportunities, specialist delays, and system constraints.
⍰ While disease progression was the perceived primary cause, IBD admissions are also perceived to be driven by modifiable systemic failures, specifically waiting times, resource gaps, and navigation hurdles influenced by social determinants.

**How this study might affect research, practice or policy:**

⍰ The findings advocate for and inform prospective service development interventions for improved rapid-access flare pathways and the strengthening of patient education to improve service navigation.
⍰ There is a clear need for significant investment in workforce and infrastructure, specifically regarding infusion capacity and specialist staffing, to prevent missed care escalation.

## Introduction

Inflammatory Bowel Diseases (IBDs) are chronic, relapsing conditions of the gastrointestinal tract that require lifelong treatment to manage flares of disease, which can result in unplanned admissions (1–3). Unplanned admissions in IBD are common as a result of severe flares and when hospital treatment becomes necessary (4–6). Whilst the clinical predictors of IBD admissions are well-researched (7–9), research specifically exploring healthcare delivery factors and how avoidable IBD admissions are understood by healthcare professionals delivering IBD care, is limited.

At current, there is limited research exploring healthcare issues in preventing unplanned IBD admissions, including prioritisation for future healthcare improvement. Previous evidence indicates that inequalities in access to care, including lack of appropriate outpatient management, are associated with IBD complications resulting in unplanned hospital admissions (10,11). Furthermore, poor quality of care appears to be leading to increased unplanned admissions or attendances to emergency departments (12), suggesting that improving care quality may prevent some unplanned IBD admissions.

To understand this complex problem and inform health service interventions to reduce unplanned IBD admissions, it is necessary to explore the perspectives of the individuals delivering healthcare (13). IBD clinicians, including gastroenterologists and nurses, attempt to reduce unplanned hospitalisations (4,11,12). Gathering their perspectives, including perceived barriers or enablers, is important to inform future interventions to reduce unplanned IBD admissions (14). Challenges relating to healthcare access have previously been explored using the Candidacy Framework, with seven interconnected constructs across individual, interpersonal, and system level influences across the healthcare journey (15–17). To our knowledge, there is no existing research exploring clinician perspectives and experiences of unplanned admissions in IBD

The aim of this study was to explore healthcare professionals’ experiences of avoidable admissions in IBD. Specific research questions were:

⍰ How do IBD healthcare professionals rank the frequency of different types of IBD admissions and causes for all unplanned IBD admissions?
⍰ How do IBD healthcare professionals define avoidable and preventable unplanned IBD admissions?
⍰ What are the perceived barriers and enablers for preventing unplanned IBD admissions?

## Materials and methods

Methods are reported using The Checklist for Reporting Results of Internet E-Surveys (CHERRIES) (18) and the Consolidated Criteria for Reporting Qualitative Research (COREQ) (19). To describe methods integration, the GRAMMS checklist for reporting of mixed-methods research in health services research is used (20) (see supplementary data A for all checklists).

### Design

A mixed methods explanatory sequential design (21) used: 1) quantitative and qualitative survey data collection; and, 2) qualitative interviews (Figure 1). Integration was built with survey outputs informing the questions and prompts of the interview guide. This approach enabled the interview data to explain survey findings. Use of a survey and interviews enabled this issue to be explored both quantitatively and qualitatively.

**Figure 1.** Flow chart of explanatory sequential mixed method design

### Setting

Experiences of multidisciplinary (MDT) IBD healthcare professionals were captured across the UK, and internationally. All participants were required to be working within a healthcare setting supporting PwIBD.

### Participants and recruitment

Eligible participants were registered IBD healthcare professionals (gastroenterologists, nurses, surgeons) and allied health professionals (pharmacists, dietitians).

Survey recruitment used opportunistic and snowball sampling over 6 months (December 2023-May 2024). Emails were distributed across UK National Health Service (NHS) IBD teams through established research links and circulated by UK IBD charity and health professional networks. The survey was advertised online via the research team ‘X’ accounts, tagging relevant IBD organisations. Sixty physical flyers were distributed at UK and European IBD professional events. Participants received no incentives.

Qualitative interview recruitment occurred via the survey over 3 months (June-August 2024). Survey participants were invited to participate in follow-up telephone calls. Those responding “yes”, and providing a contact email address, received participant information emails.

### Survey and interview guide development

Due to the absence of a pre-existing survey on IBD unplanned admissions, we constructed a new survey. Domains were selected based on existing literature (10) and collaboration with UK IBD clinicians.

An initial survey was developed and piloted for face validity, utility and feasibility using a modified QQ10 questionnaire (22) with nine UK IBD clinicians. Respondents to the modified QQ10 pilot questionnaire responded using Likert scales for statements such as “*The questionnaire included all the aspects of IBD care relevant to my practice as a clinician*”, “*the questionnaire was too long*”. Minor amendments were made following the pilot.

The final survey (supplementary B) consisted of 19 questions across 7 sections including: yes/no questions, open-questions, ranking items (scores weighting to a total of 100) and Likert scales (ranging from “never” to “all the time”) for understanding barriers. Clinicians were asked to rank four admission concepts (preventable, avoidable, unnecessary, inappropriate) by how frequently they observed each in practice. For definitions, a brief non-IBD specific definition from the literature was presented. Participants were asked if avoidable or preventable were separate terms and to provide definitions if so.

The survey was accessible on a computer or smartphone. The order of ranked question items was randomised to minimise bias. Once started, the survey remained open for two weeks allowing answer review. Question completion was required to move to subsequent sections.

The interview guide (supplementary C) was developed from survey responses and existing literature. Interviews explored topic importance, definitions and examples of avoidable and preventable IBD admissions, and perceived barriers and enablers to preventing admissions. The guide was developed with IBD clinicians (AJL, ML) during research meetings and designed to last approximately 30 minutes.

### Data collection

All data collection was remote. Survey responses were collected using Qualtrics. To protect anonymity, no personal data was collected. Participants created a unique ID code using the last two letters of their postcode and telephone number. Email addresses for interview registration were stored separately from survey data. Qualtrics automatically quality-checked through speed assessment, metadata, reCAPTCHA bot detection and duplicate detection. All responses passed quality checks. No minimum completion time was set.

A predetermined 60% completion threshold was set for final analysis inclusion. Including partial responses enables perspectives from those unable to fully engage due to time constraints and role demands. The 60% threshold was deemed sufficient given anticipated in-depth qualitative interview data.

Semi-structured telephone and GoogleMeet interviews followed a topic guide, conducted by one female PhD researcher (RH). Only participants and the researcher were present. The researcher had five years semi-structured interview experience with healthcare professionals and masters-level education. No prior participant relationships existed. Interview duration ranged 23-42 minutes. All interviews were audio recorded with participant consent. The researcher typed interview notes into a reflexive log during and after interviews.

### Analysis

Survey results were analysed using descriptive statistics. Likert responses concerning barriers to preventing admissions were transformed to numerical values (e.g., “Never”= 0, “All the time” = 7). Friedman and Wilcoxon signed rank tests assessed significant differences between ranking responses (5% significance level).

The qualitative dataset was analysed using Nvivo 14, consisting of free-text survey responses on admission definitions, combined with semi-structured interview data. This combined dataset was analysed using reflective thematic analysis (23,24). The analysis was led by one researcher (RH) with support of a second (FS). Both researchers read the transcripts and discussed themes across the dataset during meetings. Inductive coding of admission definitions was undertaken, and codes were organised into themes.

For barriers and enablers to preventing IBD admissions, reflexive thematic analysis was conducted, using the framework method (24,25). The Candidacy Framework guided analysis of access issues and barriers. Codes were organised into an analytical framework using the seven constructs of the Candidacy Framework (15–17) whilst remaining open to other codes identified inductively, beyond these constructs. Using Nvivo 14, data was charted using framework matrices and coding trees were created to explore the code clusters within the analytical framework. Case summaries were added to the chartered framework matrices summarising key messages (25). Themes were identified by code cluster analysis across the framework. A summary of findings was shared with consenting participants.

### Ethical considerations

The study received ethical approval from the Health and Care Research Support Centre, Wales Research Ethics Committee 3 (REC reference 21/WA/0264).

## Results

Eighty respondents (55.2%) met the 60% completion threshold and comprised the analysis sample for all subsequent survey results. Thirteen clinicians participated in interviews; 14 did not respond to invitations (Table 1). Nearly half of survey responses (36/80) and interviews (7/13) were with gastroenterologists. Participants varied in IBD care experience, from clinicians in training (15% of survey respondents) to those with 20+ years’ experience (21.25%). Most participants worked in tertiary IBD care (71.25% of respondents, 100% of interviews).

**Table 1.**
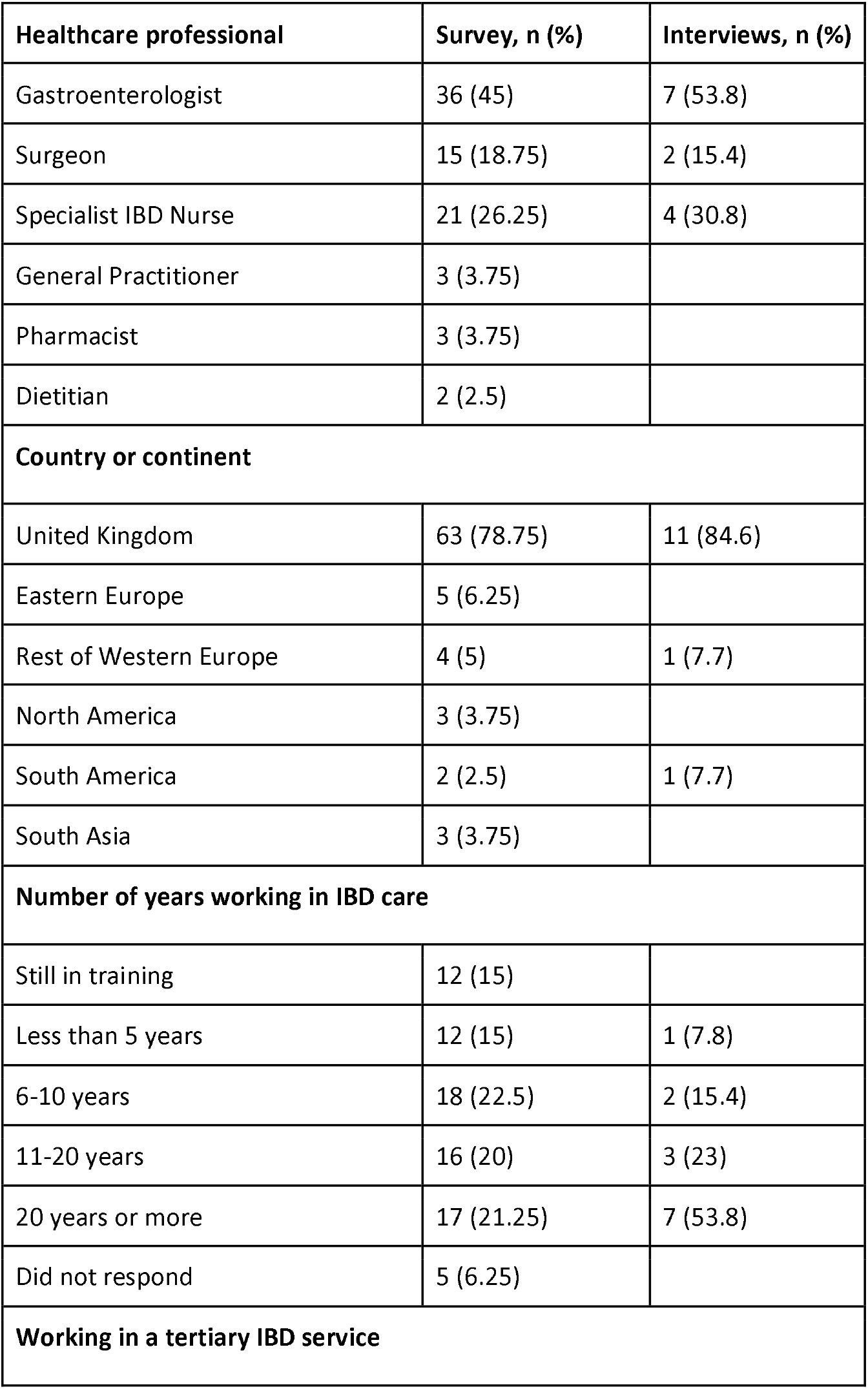
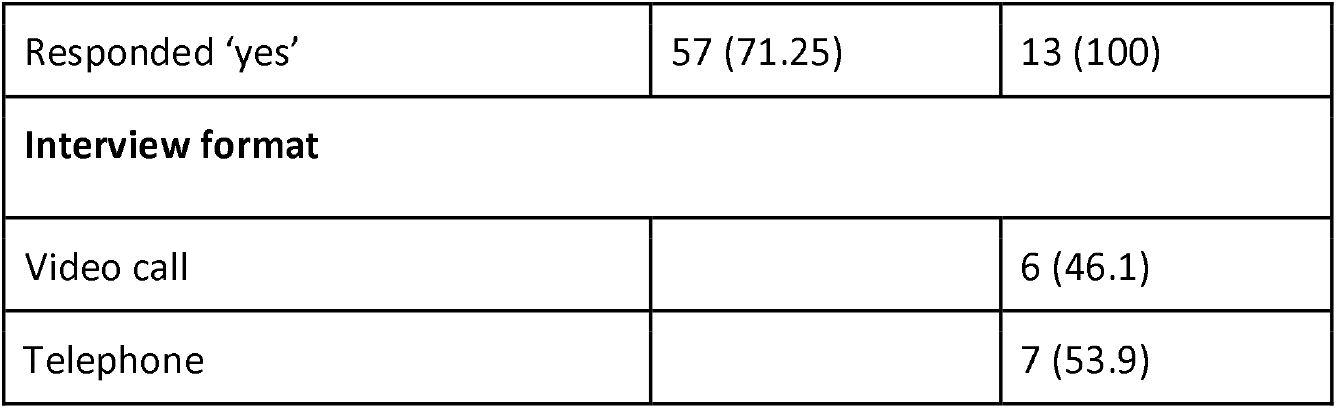
Participant demographics.

| **Healthcare professional** | **Survey, n (%)** | **Interviews, n (%)** |
| --- | --- | --- |
| Gastroenterologist | 36 (45) | 7 (53.8) |
| Surgeon | 15 (18.75) | 2 (15.4) |
| Specialist IBD Nurse | 21 (26.25) | 4 (30.8) |
| General Practitioner | 3 (3.75) |  |
| Pharmacist | 3 (3.75) |  |
| Dietitian | 2 (2.5) |  |
| **Country or continent** |  |  |
| United Kingdom | 63 (78.75) | 11 (84.6) |
| Eastern Europe | 5 (6.25) |  |
| Rest of Western Europe | 4 (5) | 1 (7.7) |
| North America | 3 (3.75) |  |
| South America | 2 (2.5) | 1 (7.7) |
| South Asia | 3 (3.75) |  |
| **Number of years working in IBD care** |  |  |
| Still in training | 12 (15) |  |
| Less than 5 years | 12 (15) | 1 (7.8) |
| 6-10 years | 18 (22.5) | 2 (15.4) |
| 11-20 years | 16 (20) | 3 (23) |
| 20 years or more | 17 (21.25) | 7 (53.8) |
| Did not respond | 5 (6.25) |  |
| **Working in a tertiary IBD service** |  |  |
| Responded 'yes' | 57 (71.25) | 13 (100) |
| **Interview format** |  |  |
| Video call |  | 6 (46.1) |
| Telephone |  | 7 (53.9) |

### Quantitative results: Survey

#### Ranking types of unplanned IBD admissions

The terms “preventable” and “avoidable” were the most favoured terms with only 5/80 clinicians ranking “inappropriate” first (Table 2). Most clinicians (57/80, 71.25%) considered avoidable and preventable admissions the same. Seventeen (21%) felt an important type of IBD admission was missing, including “necessary, unavoidable” and “urgent admission for optimisation”.

**Table 2.** Ranking of observed IBD admission concepts.

| **Admission concept** | **Overall ranking (n, % ranked first) across 80 survey responses** | **Mean rank** |
| --- | --- | --- |
| Preventable IBD admissions | 1<sup>st</sup> (46, 57.5%) | 1.62/4 |
| Avoidable IBD admissions | 2<sup>nd</sup> (22, 28%) | 2.02/4 |
| Unnecessary IBD admissions | 3<sup>rd</sup> (7, 9%) | 2.96/4 |
| Inappropriate IBD admissions | 4<sup>th</sup> (5, 6%) | 3.39/4 |
| *Across the four admission types, overall comparisons of rankings were statistically different ($p < 0.001$ Friedman test)* |  |  |

#### Reasons for all unplanned IBD admissions

For all unplanned IBD admissions, ranking of reasons (Table 3) were significantly different for all factors (*p* <0.001). Clinicians ranked unpreventable disease progression most important (MR 32.20/100) (*p* <0.05), followed by missed opportunities for earlier intervention in outpatient/community care (MR 22.39/100) and patient access to care issues (MR 19.52/100).

**Table 3.** Clinicians ranked perceived reasons for all IBD admissions.

| Ranked factor for all emergency IBD admissions |  |  | Pairwise comparisons between rankings |  |  |  |
| --- | --- | --- | --- | --- | --- | --- |
|  | Mean overall ranking | Median overall ranking | Opportunity for earlier intervention | Patient access | Patient behaviour | Clinician decision making |
|  |  |  | *Adj. p* | *Adj. p* | *Adj. p* | *Adj. p* |
| An unpreventable progression of disease requiring in-patient intervention | 32.2 | 27 | 0.40 | 0.01* | <0.001*** | <0.001*** |
| An opportunity for earlier intervention in outpatient care or the community was missed | 22.39 | 19 |  | 1 | 0.16 | <0.001*** |
| Patient access to IBD services | 19.52 | 19 |  |  | 1 | <0.001*** |
| Patient behaviour, knowledge and beliefs about illness | 15.1 | 15 | 0.16 |  |  | 0.001*** |
| Clinician decision making error | 5.5 | 5 |  |  |  |  |
| Other (please specify) | 1.0 | 0 | n=2 responses<br>1. No reason provided<br>2. "Social, hierarchy factors" |  |  |  |

Across all factors, overall comparisons of rankings were statistically significant ( $p < 0.001$ ), using the Friedman test.
Significant differences between two ranked factors were indicated using Wilcoxon Signed Rank Tests \* $p < 0.05$ , \*\*\* $p < 0.001$ . Medians & adjusted $p$ values included due to non-parametric data.

#### Barriers to preventing IBD admissions

Health service capacity and resource constraints were ranked highest (long appointment waiting times, availability of appointments and limited staff resource - 3.56-3.0/6.0) (figure 2). While ranked lower, participants recognised potentially modifiable structural barriers including lack of integration within community and outpatient services IT and referral system issues, and uncertainty in identifying patients needing early intervention (MR 1.79/6).

**Figure 2.** Multidisciplinary IBD professionals ranking of barriers to preventing IBD admissions

Sixteen (24%) participants identified additional barriers including health system capacity constraints and modifiable surgical care access issues, diagnosis/assessment/treatment issues, psychosocial circumstances of PwIBD, lack of patient support and education (Supplementary file D).

### Qualitative results: Survey and interviews

#### Definitions of avoidable and preventable IBD admissions

Three overlapping themes summarise that clinicians perceived avoidable and preventable admissions result from organisation and access issues (Figure 3; Supplementary file E): (1) Missed opportunities in outpatient or primary care, (2) Critical delays in specialised care, and (3) System constraints blocking timely action.

**Figure 3.** Diagram of clinicians’ definitions of avoidable and preventable IBD admissions

However, clinicians emphasised that *“it needs to be clear that not all admissions for IBD are avoidable”* (Specialist IBD Nurse, UK, interview) and that timely admissions are also necessary for good IBD care: *“bringing somebody in at the right time is the right thing to do”* (IBD Nurse Specialist, UK, interview). Timely admissions were not viewed as care failures.

### Barriers and enablers to preventing IBD admissions

#### Theme 1: Navigation and permeability of IBD services

Barriers and enablers to prevent IBD admissions impacting patient navigation (awareness and practical resources needed to access services) and permeability (organisation and ease of using services) are summarised using three sub-themes.

#### Inadequate system pathways and organisation

The organisation of local IBD pathways were perceived inadequate, therefore delaying IBD care. Lack of clear service organisation resulted in PwIBD experiencing navigation barriers, making preventing admissions for clinicians difficult.

Despite acknowledging that many PwIBD may present through primary care, clinicians highlighted, *“we don’t encourage patients to see their GP because the default position is to put them on steroids without any comprehensive assessment “* (Gastroenterologist, UK). Inadequate surgical referral agreements contributed to IBD admissions:

> *“We do have a surgical assessment unit… but again, our pathways to that aren’t always agreeable… there are always those that don’t meet that criteria and maybe, drop through the net a little bit*.*”*
>
> (Clinical Nurse Specialist, UK)

Clinicians described a healthcare system lacking ‘ability or resilience’, with care characterised as ‘fragmented’. Bottlenecks within the ‘conventional clinical care model’ - including the need for multiple hospital visits for testing and referrals - were seen as creating barriers to earlier intervention.:

> *“We are aware of people that are struggling to come to hospital to have multiple tests done and if you then have these patients flaring and they’re not being managed properly*.*”* (Gastroenterologist, UK)

Bottlenecks were perceived in reviewing new flares, treatment escalation, investigations, and MDT processes, delaying vital care. Clear flare pathways were perceived necessary for early intervention. Some clinicians shared positive examples:

> *“Now we have a rapid pathway so patients, if they have perianal Crohn’s they can contact the IBD nurse specialist who then arranges direct admission to hospital through a dedicated clinical fellow. Have their operation that day, and MRI within a week and with the surgeon within two weeks”* (Colorectal Surgeon, UK)

To prevent admissions, clinicians perceived reorganising services as essential, especially for flare clinics. Suggested improvements included increased administrative support, use of automatic systems, increasing available urgent appointments, and follow-up appointments.

#### Knowledge and accessing care

Lack of awareness about symptoms and when or how to contact healthcare was a common perceived barrier for navigating services (and *identification of candidacy)*. Negative previous care experiences were thought to impact how PwIBD navigate services later, delaying contact with IBD services and making preventing admissions more difficult:

> *“if patients don’t have knowledge or they’re not educated and they don’t know how to access the service, or they’re not told when to seek advice. Then absolutely that’s going to increase the likelihood of repeated hospital admissions*.*”* (IBD Nurse Specialist, UK)

Wider social barriers impacted how PwIBD navigate services. Clinicians discussed difficulties for people living in social deprivation, rural areas, or having lower health literacy. Transport barriers were linked with multiple hospital visits:

> *“I think at least here in […], some patients are not well educated in all fields so for them to understand a really complex disease”* (Gastroenterologist, South America)

This was perceived for PwIBD experiencing mental health difficulties, especially during disease flares:

> *“I think that is a bit of a problem for patients to find that number when they’re in that crisis or in that situation…They, it all gets very entangled and how to help is really difficult*.*”* (Specialist IBD Nurse, UK)

To overcome these issues and avoid admissions, patient education was perceived as important for: empowering PwIBD to contact services, to navigate healthcare, to manage expectations, and promote symptom knowledge. Signposting by IBD teams, emergency services, and charitable organisations supported navigation:

> *“I think much better education so patients understand their symptoms better, when to contact us… I think if we can get patients confident in us that they would then access us even sooner*.*”* (Gastroenterologist, UK)

##### Delays to care

Within and outside the UK, delayed care at multiple access points were perceived as a barrier to admission avoidance. This started with delays in diagnosis and referrals into specialist IBD services from primary care, resulting in admissions:

> *“I think they [primary care doctors] are not referring, they don’t refer to our hospital… that’s of course why the patients have more hospitalisations*.*”* (Gastroenterologist, rest of Western Europe)
>
> Delays were also discussed in PwIBD undergoing colonoscopies, advancing to biologics, accessing planned surgical care, and follow-up appointments. Delays in advancing treatments for IBD were highlighted in both the UK and international healthcare context. In a European hospital, they were the only centre providing biologics treatment leading to long waits. A UK clinician discussed overlapping issues:
>
> *“We are struggling to get people into these biologics. We had 150 patients waiting for biological therapy and a lot of these patients are getting admitted because we don’t have the capacity…*.*”* (Gastroenterologist, UK)

#### Theme 2: Healthcare professional adjudications

Barriers and enablers to prevent IBD admissions relating to professional adjudications (the judgements and decisions which influence subsequent access) of consultants, nurses and primary care providers were discussed across all participant interviews, summarised by two sub-themes.

#### Provider skills and confidence

For some, lacking confidence in treatment decisions amongst registrar doctors, primary care, emergency call handlers, and gastroenterologists with less experience was a perceived barrier. As advanced treatment options evolved, clinicians highlighted the need to “…*educate all the time because…They need to be able to recognise the signs and symptoms and be familiar with the novelty of the treatment so they can act*.*”* (Gastroenterologist, rest of Western Europe). This might not be possible in all cases:

> *“*…*physicians, doctors, not being familiar with the drugs, not being sure how to use them… not sure when to sequence advanced therapies… I think it’s often very difficult in the current climate to know where to go next”* (Gastroenterologist, UK)

To support treatment decisions and prevent IBD admissions, openness of communication, *“close MDT working”* (Nurse Consultant, UK), and upskilling nursing teams making complex decisions on IBD advice lines was perceived as important. Moreover, *“raising awareness at the GP level”* (IBD Clinical Nurse Specialist, UK) was perceived as important:

> *“I would say the referral to the patient. More proactive care from primary care and the internal medicine doctors… more education about the symptoms and about the steps they need to take”*
>
> (Gastroenterologist, rest of Western Europe)

#### Relational and operational obstacles

Prompt escalation of treatment was perceived essential for reducing admissions. However, system constraints, lack of time and clinician continuity negatively impacted prompt treatment decision-making: *“Our clinics don’t have the capacity to see patients in a manner that we would like to in terms of the time that we spend with them. So we see them very quickly, obviously putting us under a small degree of risk of missing things*.*”* (Gastroenterologist, UK)

Relational aspects were discussed by gastroenterologists and nurses, who shared challenges in making telephone assessments. One consultant perceived the use of interpreters in clinics as restricting clear messaging to patients. Operational challenges of making advice line decisions impacted referral decisions:

> *“*…*there’s only so much bridging you can do [on the helpline]. I requested a next available appointment today and I just got the email straight back to say for the consultant the next available appointment is in two months’ time…it stops with us on the helpline, there’s almost no point requesting*.*”* (IBD Specialist Nurse, UK)

In some services, clinicians experienced communication challenges between gastroenterology and surgical departments. Lack of safety netting processes worsened these issues, making preventing admissions more difficult:

> *“There is not always safety netting to ensure that we have checked responses to treatment…we may have initiated a treatment that is then not checked that they are actually responding to it*.*”* (IBD Clinical Nurse Specialist, UK)

### Theme 3: Local conditions and structural barriers

Local conditions (structural, societal and relational aspects) were perceived across all interviews as barriers to avoiding IBD admissions, summarised across three sub-themes.

#### Inadequate resource

Consistently, insufficient availability of local healthcare resources was perceived as a barrier. In the UK, this was *“a massive issue across the country”* (IBD Nurse Specialist, UK) and increasing IBD prevalence was perceived to compound these issues. Significant gaps included *“very few consultant gastroenterologists”* (Gastroenterologist, UK) and nurses, plus limited outpatient, infusion, inpatient and imaging capacity:

> *“The big problem we have is issues getting patients onto our infusion unit because we don’t have the capacity in our infusion unit for the number of patients that we have. Patients are waiting six to eight weeks plus to get their first dose so some patients are being admitted in order to give them their first doses as an acute admission”* (IBD Nurse Specialist, UK)

Some issues were put down to broader NHS challenges. However, across the UK and international view, a lack of approvals for accessing medications was discussed, commonly due to costs, *“the drugs are expensive, it’s the cost of the drug that is the main issue”* (Gastroenterologist, rest of Western Europe):

> *“They say it’s expensive and there doesn’t seem to be any joined up thinking that you know this delay is an expense in itself… I don’t think that’s a problem unique to our trust*” (Specialist IBD Nurse, UK).

#### Local IBD political context

Broader social and political barriers were outlined including how current services do not prioritise IBD funding, *“there’s no political power in IBD like that*.*”* (Gastroenterologist, UK). Generating discussions with healthcare management and decision-makers was perceived as challenging, “*But it just is very difficult to get IBD services on the trust in general”* (Nurse Consultant, UK).

Clinicians highlighted the competitiveness for IBD funding, especially compared to cancer services. Asking for more resource was challenging:

> *“It can be quite tricky to say that we need more staff to deal with that [the helpline], when they may just look at well, you’re going to have increasing staff costs for no increasing income”* (Gastroenterologist, UK)

Better understanding of how to prevent or avoid IBD admissions was viewed as a leverage point to increase awareness of managers, to focus minds and support adequate resourcing:

*“Because there’s some managers and others may say well that’s an acute admission it’s going to happen anyway. But I think we need to highlight as an IBD community that these are avoidable, a proportion are avoidable and preventable if we had services in place to treat our patients or treat them more promptly to avoid this*.*”* (Gastroenterologist, UK, interview)

##### Role of interpersonal relationships

Building positive relationships with PwIBD was perceived as important for reducing IBD admissions. In South America, a clinician highlighted the importance of “*Knowing the patient and knowing the history”*. Strengthening interpersonal relationships were viewed to help PwIBD feel empowered and confident to access IBD services:

> *“And actually part of our job is to try to form relationships with those patients, because those are the ones that will probably come in and are going to be admitted and that come in when they don’t have to come in. Either because they are not accessing services at all. Or because they are accessing services all the time*.*”* (IBD ACP, UK)

## Discussion

This study explored some clinicians’ perspectives on avoidable and preventable IBD admissions. Findings highlight systemic healthcare access barriers as drivers, consistent with existing literature (10,26). Clinicians perceive avoidable IBD admissions as resulting from cumulative system-level challenges rather than isolated failures, aligning with broader research on hospital admission factors (27,28).

Survey data revealed that “preventable admissions” was the most frequently observed terminology, although most participants (71.25%) used the terms “avoidable” and “preventable” interchangeably. Clinicians ranked systemic access issues as most critical causes, including long waiting times (MR 3.56/6) and poor appointment availability (MR 3.41/6). Interviews corroborated these findings, explaining that delays stem from unclear care pathways, overwhelmed clinics, and patients’ uncertainty in navigating services. These access barriers are well-documented in UK national reports (11,29).

However, data highlighted that many admissions are not preventable due to disease progression (mean rank of 32.2/100 and median of 27/100). Clinicians emphasised timely, appropriate hospitalisation remains vital to IBD care (30). Rather than attributing blame, efforts should focus on patient-centred approaches enhancing service responsiveness (31).

Clinicians defined avoidable admissions as those resulting from missed opportunities in primary or outpatient care, critical delays in accessing specialist services, or resource constraints impeding timely action. Interview examples included poor care escalation, fragmented services, and limited healthcare capacity, factors potentially contributing to IBD diagnostic delays (32). Addressing these issues requires patient-centred approaches during acute flares, incorporating mental health support, enhanced education, and improved risk communication (33).

The organisation and ease with which PwIBD were perceived to navigate healthcare presented many barriers, including appointment delays, diagnostic or infusion service bottlenecks, and poor primary-secondary care integration. These findings reinforce the need for better-integrated approaches to deliver early and personalised IBD care (34,35). Relational challenges were identified, including lack of continuity, insufficient clinician confidence in prescribing advanced therapies, and MDT inefficiencies, affecting appropriate escalation.

Qualitative interviews highlighted how social determinants affect PwIBD ‘candidacy’ for care. Clinicians described how patients with lower health literacy, language barriers, poor mental health, socioeconomic constraints, or rural residency face additional hurdles in navigating complex IBD care pathways. These findings coincide with reviews on IBD care inequalities (10), and underscore the need for equity-focused interventions. The Candidacy Framework was valuable for interpreting this interplay between patient circumstances, provider decisions, and systemic constraints (15). Thematic mapping demonstrated how access is impacted throughout the patient journey and reinforced the complexity of preventing unplanned admissions. While constructs—like “identification of candidacy” and “offers and resistance to services”—were less prominent, they emerged in discussions about treatment hesitancy and symptom awareness in newly diagnosed patients, warranting further research exploring patient perspectives.

### Implications of findings

Findings suggest several implications for improving IBD care. Services should develop clearly defined rapid-access pathways for flaring patients, supported by research modelling local service needs. Patient navigation and education must be strengthened, through early education at diagnosis, symptom recognition, and clear signposting during flares. Significant investment in the IBD workforce and infrastructure is paramount, including clinic and infusion capacity, specialist staffing, and advanced therapies to reduce costly admissions and missed care escalation opportunities. Further research is essential to understand how social and health inequities drive unplanned admissions and to develop targeted interventions.

### Strengths and Limitations

The main strength of this study is its mixed-methods design. The survey provided quantitative breadth across IBD professionals, internationally, while the interviews added depth and contextual richness. The Candidacy Framework (15) facilitated a structured understanding of barriers and enablers across multiple healthcare access domains.

Several limitations should be considered. Most participants were UK-based, limiting transferability. Survey completion rates were suboptimal, with just over half completing more than 60% of questions. Non-probability sampling contributed to unequal clinician group representation. Primary and emergency care perspectives were underrepresented in interviews, despite their critical roles in unplanned admissions. The study does not capture the experiences of PwIBD, a gap requiring further research.

### Conclusions

Clinicians in this study perceived avoidable IBD admissions as stemming from missed opportunities, delays in specialist care, and inadequate resources. Preventing these admissions requires a multifaceted approach that addresses organisational, knowledge, and interpersonal barriers. To be effective, services must be adequately resourced, flare pathways must be clearly defined, and patients must have the knowledge and access needed to receive timely care.

## Data Availability

All data in the present work are contained in the manuscript

## Financial disclosures

Funding from The University of Sheffield PhD Research Scholarships is acknowledged in supporting RLH to conduct this research.

## Conflicts of interest

AJL has acted as a speaker or consultant for Takeda, BMS, Sandoz, JNJ, Celltrion and Medtronic.

## Data availability

The data underlying this article are available in Open Science Framework (link: https://osf.io/mfxde/), in the article and in its online supplemental material provided.

## Patient and public involvement

Patients were involved in the conceptualisation and oversight of this research project via the AWARE-IBD patient panel.

## Authors contribution

RLH is a PhD student who led the conceptualisation, design, data collection, analysis and manuscript writing. DH, AJL and ML supervised the project, contributing to study design and manuscript writing. FS contributed to the analysis, interpretation of results and manuscript writing.

## Notes

### Author Declarations

The study received ethical approval from from the Health and Care Research Support Centre, Wales Research Ethics Committee 3 (REC reference 21/WA/0264)

### Summary of Updates

Study title updated Clarification of study methodology updated in the abstract and methods along with integration of survey/interview results.

