## Supplementary data for "“*We need to highlight as a community that these are avoidable*”: Exploring clinician perspectives on healthcare access and avoidable admissions in inflammatory bowel disease" (from another version of this preprint: version 1, 2025-09-14)

**Supplementary A: Reporting checklists**

*GRAMMS - O'Cathain A, Murphy E, Nicholl J. The quality of mixed methods studies in health services research. J Health Serv Res Policy. 2008;13(2):92-98*

| **Reporting Item** | **Where in manuscript** |
| --- | --- |
| (1) Describe the justification for using a mixed methods approach to the research question | Methods |
| (2) Describe the design in terms of the purpose, priority and sequence of methods | Methods, Figure 1 |
| (3) Describe each method in terms of sampling, data collection and analysis | Methods |
| (4) Describe where integration has occurred, how it has occurred and who has participated in it | Methods, Figure 1 |
| (5) Describe any limitation of one method associated with the present of the other method | Discussion - strengths and limitations |
| (6) Describe any insights gained from mixing or integrating methods | Discussion |

[***LINK TO*** *COREQ (COnsolidated criteria for REporting Qualitative research) Checklist*](https://docs.google.com/document/d/1fpI41liWXiJYH0-yWbgKtKU5jnaQfOoj8K1nX4-bbWY/edit?tab=t.0)

*Checklist for Reporting Results of Internet E-Surveys (CHERRIES)*

| **Checklist Item** | **Explanation** | **Page Number** |
| --- | --- | --- |
| Describe survey design | Describe target population, sample frame. Is the sample a convenience sample? (In “open” surveys this is most likely.) | 7 |
| Institutional Review Board (IRB) approval | Mention whether the study has been approved by an IRB. | 10 |
| Informed consent | Describe the informed consent process. Where were the participants told the length of time of the survey, which data were stored and where and for how long, who the investigator was, and the purpose of the study? | 6-7 |
| Data protection | If any personal information was collected or stored, describe what mechanisms were used to protect unauthorized access. | 8 |
| Development and testing | State how the survey was developed, including whether the usability and technical functionality of the electronic questionnaire had been tested before fielding the questionnaire. | 7-8 |
| Open survey versus closed survey | An “open survey” is a survey open for each visitor of a site, while a closed survey is only open to a sample which the investigator knows (password-protected survey). | 8 |
| Contact mode | Indicate whether or not the initial contact with the potential participants was made on the Internet. (Investigators may also send out questionnaires by mail and allow for Web-based data entry.) | 8 |
| Advertising the survey | How/where was the survey announced or advertised? Some examples are offline media (newspapers), or online (mailing lists – If yes, which ones?) or banner ads (Where were these banner ads posted and what did they look like?). It is important to know the wording of the announcement as it will heavily influence who chooses to participate. Ideally the survey announcement should be published as an appendix. | 6-7 |
| Web/E-mail | State the type of e-survey (eg, one posted on a Web site, or one sent out through e-mail). If it is an e-mail survey, were the responses entered manually into a database, or was there an automatic method for capturing responses? | 7-8 |
| Context | Describe the Web site (for mailing list/newsgroup) in which the survey was posted. What is the Web site about, who is visiting it, what are visitors normally looking for? Discuss to what degree the content of the Web site could pre-select the sample or influence the results. For example, a survey about vaccination on a anti-immunization Web site will have different results from a Web survey conducted on a government Web site | 7 |
| Mandatory/voluntary | Was it a mandatory survey to be filled in by every visitor who wanted to enter the Web site, or was it a voluntary survey? | 8 |
| Incentives | Were any incentives offered (eg, monetary, prizes, or non-monetary incentives such as an offer to provide the survey results)? | 7 |
| Time/Date | In what timeframe were the data collected? | 7 |
| Randomization of items or questionnaires | To prevent biases items can be randomized or alternated. | 7-8 |
| Adaptive questioning | Use adaptive questioning (certain items, or only conditionally displayed based on responses to other items) to reduce number and complexity of the questions. | 7-8 |
| Number of Items | What was the number of questionnaire items per page? The number of items is an important factor for the completion rate. | 7 |
| Number of screens (pages) | Over how many pages was the questionnaire distributed? The number of items is an important factor for the completion rate. | 7 |
| Completeness check | It is technically possible to do consistency or completeness checks before the questionnaire is submitted. Was this done, and if “yes”, how (usually JAVAScript)? An alternative is to check for completeness after the questionnaire has been submitted (and highlight mandatory items). If this has been done, it should be reported. All items should provide a non-response option such as “not applicable” or “rather not say”, and selection of one response option should be enforced. | 8 |
| Review step | State whether respondents were able to review and change their answers (eg, through a Back button or a Review step which displays a summary of the responses and asks the respondents if they are correct). | 8 |
| Unique site visitor | If you provide view rates or participation rates, you need to define how you determined a unique visitor. There are different techniques available, based on IP addresses or cookies or both. | DNR |
| View rate (Ratio of unique survey visitors/unique site visitors) | Requires counting unique visitors to the first page of the survey, divided by the number of unique site visitors (not page views!). It is not unusual to have view rates of less than 0.1 % if the survey is voluntary. | DNR |
| Participation rate (Ratio of unique visitors who agreed to participate/unique first survey page visitors) | Count the unique number of people who filled in the first survey page (or agreed to participate, for example by checking a checkbox), divided by visitors who visit the first page of the survey (or the informed consents page, if present). This can also be called “recruitment” rate. | DNR |
| Completion rate (Ratio of users who finished the survey/users who agreed to participate) | The number of people submitting the last questionnaire page, divided by the number of people who agreed to participate (or submitted the first survey page). This is only relevant if there is a separate “informed consent” page or if the survey goes over several pages. This is a measure for attrition. Note that “completion” can involve leaving questionnaire items blank. This is not a measure for how completely questionnaires were filled in. (If you need a measure for this, use the word “completeness rate”.) | 11 |
| Cookies used | Indicate whether cookies were used to assign a unique user identifier to each client computer. If so, mention the page on which the cookie was set and read, and how long the cookie was valid. Were duplicate entries avoided by preventing users access to the survey twice; or were duplicate database entries having the same user ID eliminated before analysis? In the latter case, which entries were kept for analysis (eg, the first entry or the most recent)? | DNR |
| IP check | Indicate whether the IP address of the client computer was used to identify potential duplicate entries from the same user. If so, mention the period of time for which no two entries from the same IP address were allowed (eg, 24 hours). Were duplicate entries avoided by preventing users with the same IP address access to the survey twice; or were duplicate database entries having the same IP address within a given period of time eliminated before analysis? If the latter, which entries were kept for analysis (eg, the first entry or the most recent)? | 8 |
| Log file analysis | Indicate whether other techniques to analyze the log file for identification of multiple entries were used. If so, please describe. | DNR |
| Registration | In “closed” (non-open) surveys, users need to login first and it is easier to prevent duplicate entries from the same user. Describe how this was done. For example, was the survey never displayed a second time once the user had filled it in, or was the username stored together with the survey results and later eliminated? If the latter, which entries were kept for analysis (eg, the first entry or the most recent)? | N/A |
| Handling of incomplete questionnaires | Were only completed questionnaires analyzed? Were questionnaires which terminated early (where, for example, users did not go through all questionnaire pages) also analyzed? | 8 |
| Questionnaires submitted with an atypical timestamp | Some investigators may measure the time people needed to fill in a questionnaire and exclude questionnaires that were submitted too soon. Specify the timeframe that was used as a cut-off point, and describe how this point was determined. | 8 |
| Statistical correction | Indicate whether any methods such as weighting of items or propensity scores have been used to adjust for the non-representative sample; if so, please describe the methods. | DNR |

**Supplementary B: Survey**

Start of Block: Section 1: Information about survey completers

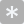

**Please create your own unique participant identification code (last 2 numbers of your phone number & last 2 letters of your postcode)**

_______________________________________________________________

**What is your current professional position/job title?**

o Gastroenterologist (1)

o General Practitioner (2)

o Surgeon (3)

o Other (4)

o Specialist IBD nurse (5)

**If other, please state your professional job title below**

________________________________________________________________

**How long have you worked with IBD patients as a consultant or specialist nurse?**

o Still in training (1)

o Less than 5 years (2)

o 6-10 years (3)

o 11-20 years (4)

o 20 years or more (5)

o Not applicable (6)

**Where in the world are you based?**

o In the UK (1)

o Outside of the UK (2)

**In what region of the UK do you currently work with IBD patients?**

o North East (1)

o North West (2)

o Yorkshire and The Humber (3)

o East Midlands (4)

o West Midlands (5)

o East of England (6)

o London (7)

o South East (8)

o South West (9)

o Scotland (10)

o Wales (11)

o Northern Ireland (12)

**In what country do you currently work with IBD patients?**

________________________________________________________________

**Does your centre or practice provide tertiary care for IBD? (receive referrals from other hospitals)**

o Yes (1)

o No (2)

o I am not sure (3)

o Not applicable (4)

**What IBD specialist MDT members do you have access to in your centre/practice?**

▢	Colorectal Surgeon (1)

▢	Radiologist (2)

▢	Psychologist (3)

▢	Dietician (4)

▢	Specialist Gastroenterologist (5)

▢	Specialist Nurse (6)

▢	Social worker (7)

▢	Physiotherapist (8)

End of Block: Section 1: Information about survey completers

Start of Block: Section 2: Consensus on terminology/definitions

There are a range of terms and definitions that are used interchangeably to describe unplanned emergency admissions across chronic health conditions.	Below we provide a list of the types of admissions we are interested in exploring further in our research: Preventable admission	Avoidable admission	Unnecessary admission	Inappropriate admission

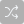

**How often do you observe or experience these types of admission in your practice? Please rank them in order.**

______ Preventable admission (1)

______ Avoidable admission (2)

______ Unnecessary admission (3)

______ Inappropriate admission (4)

**Is there an important term or type of admission in IBD missing from this list?**

o Yes (1)

o No (2)

**If you would use a different terminology or language than what has been listed in this section of the survey, please enter it here.**

________________________________________________________________

**How would you define this type of admission within the context of IBD care?**

________________________________________________________________

Our research is interested in understanding how some emergency admissions in IBD can be avoided or prevented through quality improvement and community-based preventative IBD care. **As a healthcare professional working in IBD care, do you think preventable and avoidable admissions are the same, or different?**

o They are the same (1)

o They are different (2)

**How would you define an avoidable unplanned admission within the context of IBD care?**

________________________________________________________________

**How would you define an preventable unplanned admission within the context of IBD care?**

________________________________________________________________

End of Block: Section 2: Consensus on terminology/definitions

Start of Block: Section 3: Factors

Below contains a list of a consensus on the individual and healthcare system factors associated with an IBD emergency admission. This list has been created based on engagement with stakeholders that included IBD expert clinicians and IBD patients, as well as evidence from the literature.

| 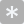 | 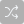 |
| --- | --- |

**Based on the IBD patients you have seen over the past year, which of these factors do you feel are most associated with emergency admissions in IBD?** Please enter the score on the right hand side of the chart. Your score will add up to 100

_______ **An unpreventable progression of disease requiring in-patient intervention** (intestinal obstruction, suspected sepsis/perianal sepsis) (1)

_______ **An opportunity for earlier intervention in outpatient care/ the community was missed that could have prevented the admission** (early escalation of steroids) (2)

_______ **Clinician decision making error** (cognitive errors, biases, catagorisation of the situation) (3)

_______ **Patient behaviour, knowledge and beliefs about illness** (knowledge to contact service during a flare) (4)

_______ **Patient access** (referral waiting times, ease of accessing IBD services) (5)

_______ **I don't know** (6)

_______ **Other (please specify)** (7)

**Please explain below what "other" factors you feel are associated with an unplanned IBD admission?**

________________________________________________________________

________________________________________________________________

________________________________________________________________

________________________________________________________________

________________________________________________________________

End of Block: Section 3: Factors

Start of Block: Section 4: Barriers

Our research is interested in understanding how some emergency admissions in IBD might possibly be prevented or avoided through changes within the health system. This section below describes some barriers identified in the literature relating to healthcare system that are associated with emergency admissions across chronic health care conditions.

A range of different factors can influence if and how an unplanned admission could be prevented. **To what extent have you experienced the following issues at your local trust?**

|  | Never (1) | Rarely (less than 10% of the time) (2) | Occasionally (about 30% of the time) (3) | Sometimes (about 50% of the time) (4) | Frequently (about 70% of the time) (5) | Usually (about 90% of the time) (6) | All the time (7) |
| --- | --- | --- | --- | --- | --- | --- | --- |
| Inadequate IT/ Referral systems (1) | o | o | o | o | o | o | o |
| Long appointment waiting times (2) | o | o | o | o | o | o | o |
| Limited availability of appointment/clinic slots (3) | o | o | o | o | o | o | o |
| Access to, and appropriate organisation of IBD helplines (4) | o | o | o | o | o | o | o |
| Limited staff resource and capacity e.g., shortages, lack of time spent with patients (5) | o | o | o | o | o | o | o |
| Uncertainty in identifying patients in need of early intervention (6) | o | o | o | o | o | o | o |
| Degree of integration with community / outpatient services (7) | o | o | o | o | o | o | o |

**Are there any other barriers you have experienced that are missing from this list?**

o Yes (1)

o No (2)

**What other barriers do you experience?**

________________________________________________________________

**To what extend do you experience this barrier in your everyday practice?**

|  | Never (1) | Rarely (less than 10% of the time) (2) | Occasionally (about 30% of the time) (3) | Sometimes (about 50% of the time) (4) | Frequently (about 70% of the time) (5) | Usually (about 90% of the time) (6) | All the time (7) |
| --- | --- | --- | --- | --- | --- | --- | --- |
| Please type barrier here (4) | o | o | o | o | o | o | o |

End of Block: Section 4: Barriers

Start of Block: Block 6

**You are about to submit your answers. Before you do so, is there anything else you would like to add about this topic?**

________________________________________________________________

End of Block: Block 6

Start of Block: Section 5: Consent for future contact

In this survey we have used terms with lots of overlapping meaning, we are interested in hearing more about what they mean for IBD management. If you would be happy to be contacted about this please leave your email address.

________________________________________________________________

Would you like to receive a copy of the results of this study?

o Yes (1)

o No (2)

Please enter your email address to be emailed a copy of the survey results

_______________________________________________________________

**Supplementary C: Semi-structured interview guide**

**1.** **Can you tell me about your professional job role, and what interested you in this study?**

Prompts:

- Do you think this research is important, why is this (topic) important?
- Do you feel this is an issue in your hospital/centre?

**2.**	**There are different concepts used to describe unplanned IBD admissions (including avoidable, preventable, unnecessary, unwarranted admissions). How important is it that we are able to distinguish between the use of these terms?**

Prompts:

- Does this have any implications on practice?
- Why is this important/not important
- Does it affect your decision making prior to or during an admission?
- Does it affect your decision making prior to introducing advanced therapies?
- Do you think using those concepts would change the way you use those drugs? Earlier?

**3.** **Our research so far has shown that some clinicians view concepts of "avoidable" and "preventable" hospital admissions as the same, whilst some view them as different. What do you think?**

Prompts:

- Why do you think this is?
- How would you define these types of admissions?
- Does improper use of these terms have implications on your practice?

**4.**	**Research suggests that inequalities in accessing healthcare, and experiences of IBD healthcare may be a driver of avoidable admissions in IBD. What do you think about this?**

Prompts:

- What does experience of care mean to you?
- What do you see as important drivers to unplanned IBD admissions?
- Do you see this as an equity of care problem?
- Do you foresee service changes closing gaps in IBD care quality?

**5.** **Research suggests that optimising care in outpatient settings may contribute to reducing admissions. What role does your healthcare setting (e.g. outpatient, inpatient, primary care) play in avoiding unplanned admissions?**

Prompts:

- What would ‘optimal’ outpatient care look like to you for an IBD patient?

**6.** **Have we missed anything that you feel is key in preventing some IBD admissions?**

Prompts:

- What are the barriers to preventing admissions?
- Are the ways you would prevent admissions different in avoidable vs preventable admissions?
- Given that we want to prevent both, are the measures you would take different? i.e. there are some measures that would prevent both?
- Why is this person/actor important? What role do they play?
- Complexity of systems, patient navigation through systems/referrals, access issues, service capacity/staffing, IT systems

**7.** **Those are all my questions. Do you have anything else you would like to add that we haven’t talked about today?**

**Supplementary D:**

*Table 4. Free-text responses of barriers to preventing unplanned IBD admissions*

| **Barrier category** | **Quotes from survey responses** | **Relevance to Candidacy** |
| --- | --- | --- |
| **Health system capacity and resource constraints** | - *"Infusion capacity leading to unacceptable delays" (IBD Advanced Nurse Practitioner, UK)* - *"Lack of acute care unit or same day emergency care in tertiary care that will review IBD patients where admission may not be necessary" (Gastroenterologist, UK)* - *"Clinical nurse specialist has identified urgent need but no consultant clinic slots" (Specialist IBD Nurse, UK)* - *"Specialist dietitian and nutritional support (staffing)"* (Surgeon, UK) - *"Constant pressure on our IBD service line, we respond effectively but its very demanding"* (Nurse Consultant, UK) | **Permeability of IBD Services** - Limited capacity (infusion and clinic slots, staffing) creates barriers to accessing IBD services  **Operating Conditions and Local Production of Candidacy** - Limitations in available local specialist IBD resources that impact service provision |
| **Surgical access issues** | - *"Poor surgical responses and support i.e. I&D, emergency surgery, long waits for elective surgery" (Specialist IBD Nurse, UK)* - *"Access to theatre lists"* (Surgeon, UK) - *"Lack of ability to get patients on urgent theatre lists"* (Surgeon, UK) - *"Lack of ambulatory emergency lists (e.g. for perianal crohn's patients)"* (Surgeon, UK) | **Permeability of IBD Services** - Difficulties accessing surgical services and theatre lists create barriers in the patient journey  **Operating Conditions and Local Production of Candidacy** - Limited surgical resources and lack of ambulatory emergency lists reflect local issues |
| **Challenges with diagnosis, clinical assessments and treatment** | - *"Lack of primary care early diagnosis" (Gastroenterologist, South America)* - *"Inadequate assessment in telephone follow up by medical teams" (Surgeon, UK)* - *"Patients being referred in on two week wait because FIT testing and then multiple people are involved causing confusion" (Specialist IBD Nurse, UK)* - *"Delays with starting patients on homecare medication" (Specialist IBD Nurse, UK)* | **Adjudications by Healthcare Professionals** - Inadequate telephone assessments reflect issues in professional judgment of disease status  **Permeability of IBD Services** - Complex referral pathways with multiple services, people and professionals create navigation barriers |
| **Psychosocial circumstances and lack of support** | - *"Social circumstances and lack of IBD psychologist support"* (IBD/Gastro Fellow, UK) - *"Admissions based on anxiety"* (IBD/Gastro Fellow, UK) - *"Underserved communities i.e. ethnicity and deprivation”* (Gastroenterologist, UK) | **Operating Conditions and Local Production of Candidacy** - Lack of IBD psychologist support reflects limitations in available specialised resources |
| **Lack of patient education** | - *"Lack of early patient education resulting in inappropriate IBD helpline use"* (IBD/Gastro Fellow, UK) | **Navigation of IBD Services -** Education impacts awareness of options like IBD helplines as first points of contact during flares |
| **Other (uncategorised)**  **Note* these barriers were also uncategorised due to lack of clarity in meaning of response* | - *"Lack of compassion for IBD patients" (Gastroenterologist, rest of Western Europe)* - *"When medicine discounts the value of other professional opinions" (Specialist IBD Nurse, UK)* |  |

**Supplementary E:**

*Table 5. Additional quotes from clinician definitions of avoidable and preventable IBD admissions*

| **Theme** | **Codes** | **Example quote, participant** |
| --- | --- | --- |
| **Missed opportunities for outpatient support, early intervention, and follow-up** | Lack of access | *“Not able to access advice and they are brought in” (Surgeon, UK, survey)*  *“Lack of access to specialist nurse or flare up treatment leading to an acute admission” (*General Practitioner, UK)  “Patients don’t have the best health access, treatment or follow-up” (Gastroenterologist, South America, survey) |
|  | Unable to contact services | *“The usual scenario that we see is patients with Ulcerative Colitis who have a little bit of blood that they start to see three or four months before, can’t get in touch with everybody, don’t get an early course of steroids to get things under control, don’t get on a course of biologics if they are moderate to severe disease and then they present with acute-severe colitis”* (Gastroenterologist, UK, interview)  “An avoidable admission is where someone had relatively moderate symptoms, tried several times to get in touch with healthcare professionals, tried many times, and ended up in A&E” (Gastroenterologist, UK, interview) |
|  | Patient education | *“And also the education of the patient. They need to be aware of the symptoms and report early and not say ok this is something that will pass. They need to know the evolution and path of the disease and recognise the early symptoms of exacerbation.”* (Gastroenterologist, *rest of Western Europe*, interview)  *“That probably was because we didn’t educate them back at the start of their journey when they were diagnosed. So I think even when we, you know it is very easy to blame patients and say they presented late but, you know I think that’s a bit of a cop out.”* (Gastroenterologist, UK, interview) |
|  | Patient-provider rapport | *“And then that’s about, that’s something that probably could have been avoided if we had better education of the patient or better rapport with the patient.”* (Specialist IBD Nurse, UK, interview) |
|  | Lack of follow up | *“Admission due to lack of follow up of the patient”* (Gastroenterologist, Egypt, survey)  “I would define this as an admission resulting from a lack of action or incorrect action taken by a member of the primary care team following contact with the patient” (General Practitioner, UK, survey) |
|  | Acting earlier | *“Admitted for severe flare up of disease without medication having been escalated first before it got to this stage of severity”* (Trainee Doctor in Gastroenterology, UK, survey)  *“...but in my mind if I have a regular check up with the patient. I can see the signs and symptoms. I can use the tools that I have like the CDI scoring system and I can see what can happen in the future and I can act earlier so that the hospitalisation will be prevented.”* (Gastroenterologist, *rest of Western Europe*, interview)  “an earlier intervention [that] might have prevented the admission” (Specialist IBD Nurse, UK, survey  “[...]Maybe developing a perianal abscess. It’s then that earlier intervention with maybe surgical intervention rather than allowing things to develop whereby they are then requiring sort of surgery to rectify things.” (IBD Nurse, UK, interview) |
|  | Dealt with via helpline/ in clinic | *“Avoidable is more in the remit of gastroenterologists. People can be managed in the community as an outpatient then that is a wasted bed if the patient comes in and wasted time for the patient.”* (Surgeon, UK, interview)  “*Patient being admitted for intravenous steroids who may have been more appropriately managed with oral therapy first (i.e. did not satisfy severity criteria in addition to clinical assessment)*” (Gastroenterologist, UK, survey) |
| **Critical delays in specialised care** | Delays in treatment escalation | *“You do think at the time that you know, had they been seen sooner that might have avoided an admission if the treatment had been started sooner. I suppose it’s waiting for tests as well potentially. So yeah, that would be the most obvious thing. Delays causing admissions.”* (Specialist IBD Nurse, UK, interview)  *“Admission that can be prevented is adequate treatment is given in time”* (Surgeon, UK, survey)  “Somebody with ileocolonic Crohn’s disease is gradually getting a little bit of obstructive symptoms, doesn’t get to healthcare, treatment isn’t modified, it is not escalated in time and eventually ends up getting admitted…” (Gastroenterologist, UK, interview) |
|  | Delays in clinical assessments/reviews | *Somebody who might not be able to get a clinic date, awaiting to get into clinic to start a new treatment erm and we’ve had a few calls to the helpline…, they’re not treatments we start on the helpline. So people will be waiting for the appointment to start that drug and then they’ll call the helpline. So, if those patients haven’t called or if they call and they’ve got so bad then sometimes they will have to be admitted”* (Specialist IBD Nurse, UK, interview)  *“Admission that can be avoided if early symptoms of exacerbation are predicted or seen on time*” (Gastroenterologist, *rest of Western Europe*)  “Timely, appropriate medical/surgical review with sound management plans in place” (Specialist IBD Nurse, UK, survey) |
|  | Delays in referrals | *“So, an avoidable admission means that there are things that could have been done either in the community or through the outpatient clinic, that haven’t happened which have then led to a patient being admitted. So whether that was an early referral to a surgeon…”* (Surgeon, UK, interview)  “An obvious thing would be somebody who might be a newly diagnosed person or waiting for a diagnosis where they have been referred to the service and they are waiting for an investigation. They get a diagnosis, but eventually just get worse and then they have to be admitted. That would be avoidable.” (Specialist IBD Nurse, UK, interview) |
| **System constraints blocking timely action** | Windows of opportunity | *“Whereas avoidable is something that happens over months, so you’ve got that window of opportunity to act over several weeks and the system isn’t geared to address that issue over a period of several weeks then eventually that avoidable admissions becomes that emergency and comes in. So, an avoidable admission allows that system to at least prepare itself and with a certain degree of resource implications, put into place a plan to prevent that avoidable from becoming an emergency.”* (Specialist IBD Nurse, UK, interview) |
|  | Available structures | *“One where the structure of ambulatory and community IBD care allows optimisation of the condition reducing the need for unplanned admission”* (Gastroenterologist, UK, survey)  “Given advice but non resources for it to happen” (Surgeon, UK, survey).  “So, an avoidable admission allows that system to at least prepare itself and with a certain degree of resource implications, put in to place a plan to prevent that avoidable from becoming an emergency” (Gastroenterologist, UK, interview) |
|  | Timely treatment | *“We had loads of them last year because we had lots of patients who we couldn’t get commenced on treatment because we had capacity issues in our infusions unit. Which were really clear erm and it was very difficult to manage. So we had lots of patients who were admitted because we couldn’t get them started on treatment in a timely fashion. That’s preventable. It's highly preventable. It’s absolutely appalling that that happened. Disgusted, you know. But that’s a very preventable admission.*” (Specialist IBD Nurse, UK, interview)  “So we had lots of patients who were admitted because we couldn’t get them started on treatment in a timely fashion. That’s preventable. It's highly preventable.” (Specialist IBD Nurse, UK, Interview) |
